## Supplemental_Methods for "Common genetic variation associated with Mendelian disease severity revealed through cryptic phenotype analysis"

August 26, 2021

As discussed in the main text, phenotypic heterogeneity among Mendelian disease cases is common. Although this within-disease variation is well recognized, the precise contributing factors (genetic variants, environmental exposures, etc) remain difficult to identify [1]. This is likely due to the limited sample sizes available when studying Mendelian disorders, and researchers have adopted several strategies to overcome this shortcoming, including the generation of pooled cohorts [2, 3], genetic analyses in model systems [4, 5], and the statistical integration of orthogonal datasets [6]. Here, we outline a new approach for detecting genetic modifiers of Mendelian disease severity that directly addresses the issue of limited sample size. It does so by expanding the set of available subjects to include individuals that have similar phenotypes but not necessarily the Mendelian disease itself.

Crucially, the success of this approach relies on two assumptions. First, a spectrum of cryptic phenotypic variation must exist within the population such that the Mendelian disease cases tend to cluster towards the most severely affected extreme. However, there should also be many other patients in the population with similar phenotypes, which are instead driven by other genetic/environmental factors. Second, our method assumes that the modifiers of disease severity are constant across the different etiologies. If both of these assumptions are valid, then the effects of Mendelian disease modifiers should be apparent in a larger subset of patients, resulting in increased power to detect associations.

The remainder of this text is structured as follows. In Section 1, we describe in detail the datasets used in our analyses. Section 2 outlines our approach to automatically detecting rare disease diagnoses and their associated symptoms within structured medical data. Finally, Section 3 describes our methods for estimating cryptic spectrums of phenotypic severity from observed symptoms. The remainder of the Methods are covered in the primary text. Supplemental Figures and Tables are included at the end of this document.

#### 1 Clinical Datasets

This section describes our processing of the clinical datasets used for latent phenotype inference and the subsequent genetic analyses (UCSF De-Identified Clinical Data Warehouse: UCSF-CDW; UK Biobank: UKBB). Neither dataset can be shared directly with third parties, as both have specific requirements against open data sharing outside of their usual application processes. Information regarding third party access to the UCSF De-Identified Clinical Data Warehouse can be found through UCSF Data Resources[1], and the application process for access to the UK Biobank is outlined on their website[2].

##### 1.1 UCSF De-Identified Clinical Data Warehouse

We performed all initial phenotypic analyses and modeling using the University of California San Francisco De-Identified Clinical Data Warehouse (UCSF-CDW)[3], a database of structured electronic health information

[1] `https://data.ucsf.edu/cdrp/research`
[2] `https://www.ukbiobank.ac.uk/register-apply/`
[3] `https://myresearch.ucsf.edu/de-identified-clinical-data-warehouse`

that is made available to UCSF researchers free-of-charge. The data was captured for use as flat text files on May 31st, 2019 and not updated thereafter. Following capture, patient demographic data (age in decades, sex, ethnicity, multiracial identifier, language, mortality status, marital status, and smoking status) was aligned to the medical encounter diagnoses documented in the form of International Classification of Disease, Tenth Revision, Clinical Modification (ICD10-CM) codes [7]. This generated demographic and diagnostic information for 1,204,212 de-identified patients who have received their healthcare within the UCSF system since 2012 (along with some migrated legacy data dating back to 1988).

To further simplify the clinical information, we constructed a binary vector for every patient indicating whether or not they had ever been assigned each ICD10-CM code (stored in a sparse format, see `ClinicalDataset` module in `vlpi` package on `https://github.com/daverblair/vlpi` for details). Note, this *at-least-one* binarization of diagnostic information was chosen to maximize the sensitivity of the encodings at the expense of specificity. Although this approach may not be ideal when conducting single-trait association studies [8, 9, 10], the majority of our analyses instead rely on capturing the correlation structure among multiple diagnostic codes and should therefore be robust against isolated false positives.

After parsing the diagnostic codes into binary vectors, we removed several classes of ICD10-CM codes from the dataset. First, we removed all codes that represent diagnoses related to external causes (ICD10-CM Chapters 19 and 20), factors affecting health status (Chapter 19), and diagnoses related to pregnancy/the perinatal period, which appeared to be inconsistently used across mother and child (Chapter 15 and Chapter 16-P09). We then removed all ICD10-CM codes identifying each rare disease of interest (see `SupplementalDataFile_1.txt` for mappings; further details provided below), storing these diagnoses as binary vectors (again using *at-least-one* binarization) for use in downstream analyses. Additionally, we found that ICD10-CM codes within the same hierarchical level as a rare disease of interest were often highly predictive of said disease even when intending to document a fairly distinct clinical entity. This suggests such codes can serve as noisy but unintentional proxies for rare disease diagnoses, which would bias some of our downstream analyses. Therefore, they were removed from the dataset as well. Finally, we removed all ICD10-CM codes with a frequency less than $10^{-5}$, as such codes offered little information at the expense of much dataset/analytical complexity. This processing generated a binary array (present/absent) of $10,483$ diagnostic terms assigned to $\approx 1.2$ million patients (subsequently denoted *UCSF-ICD10-CM*).

We further processed *UCSF-ICD10-CM* in two ways. First, we transformed the ICD10-CM codes into HPO terms using a customized mapping, whose construction is outlined in detail below (resulting dataset denoted *UCSF-HPO*). Second, we translated the ICD10-CM codes into the ICD10 terminology utilized by the UK Biobank[4], taking advantage of the fact that the UK Biobank encoding is a less granular subset of the ICD10-CM encoding (see `ICD10TranslationMap` in `vlpi.ICDUtilities` for details concerning the implementation of said mapping). This dataset will subsequently be referred to as *UCSF-ICD10-UKBB*. These processed datasets contained $1,674$ and $4,933$ diagnostic terms respectively. The *UCSF-ICD10-UKBB* dataset was also translated into HPO terms, resulting in a dataset with $1,423$ features (denoted *UCSF-HPO-UKBB*).

The UCSF dataset was used for statistical model inference and evaluation. Therefore, it was *a priori* divided into two subsets: a *training dataset* and a *testing dataset*. We wanted to ensure that the testing dataset contained positive cases for each rare disease included in our analysis, so distinct training and testing datasets were generated for each disorder. The subsets were constructed by randomly subsampling 75% of the data for training and 25% for testing while maintaining an equal ratio of rare diseases cases in each. All inference and preliminary analyses were performed using the training dataset, while the testing dataset was only used to perform our final evaluations of the cryptic phenotypes (see below). Training and testing datasets were generated using the `ClinicalDatasetSampler` class available within the `vlpi` software package: `https://github.com/daverblair/vlpi`.

#### 1.2 UK Biobank

The bulk UK Biobank (UKBB) dataset was downloaded on Jan 22nd, 2020 using the software provided by the organization[5]. Following download, the raw data file was parsed, isolating demographic variables of

[4]Data-Coding 19: `https://biobank.ndph.ox.ac.uk/showcase/coding.cgi?id=19`

[5]See `https://biobank.ctsu.ox.ac.uk/crystal/exinfo.cgi?src=accessing_data_guide` for details regarding data access.

interest and collapsing main/secondary inpatient summary diagnoses into a single data value[6] . This parsed dataset was then loaded into a `vlpi.data.ClinicalDataset` class. Finally, the ICD10 codes in the raw dataset were filtered by removing:

1. Codes identifying a rare disease of interest (see `SupplementalDataFile_1.txt` for precise mappings)
2. Codes from the disallowed set described above
3. Codes not contained within the translated UCSF dataset (i.e. *UCSF-ICD10-UKBB*)

This resulted in a version of the UK Biobank dataset (denoted *UKBB-ICD10*) whose diagnostic codes had a 1:1 mapping to those available within *UCSF-ICD10-UKBB* (4,933 diagnostic terms). This dataset was also translated into HPO terms (denoted *UKBB-HPO*), yielding a sparse, binary array with 1,423 features. Because this dataset was used for phenotype model inference, training (random 75%) and testing (random 25%) subsets were also constructed for each Mendelian disease (see above).

The processing described above was applied to the full UKBB dataset. However, subsets of the full dataset were used for the genetic analyses based on recommended best practices [11, 12]. First, the full dataset (i.e. all individuals with imputed genotype data after removing withdrawn subjects) was further filtered by removing subjects with:

1. Self-reported/genetic sex mismatch
2. Sex-chromosome aneuploidy
3. Heterozygosity/missingness outliers
4. Genotype call rate $< 97\%$
5. Kinship outlier status

This reduced the full dataset from 487,297 to 485,014 subjects (denoted *UKBB-ICD10-Full* in the remainder of the text). Next, two additional subsets of the data were constructed. The first included only Caucasian-identifying individuals whose ethnicity matched that inferred by Principal Component Analysis of the genetic relationship matrix (*UKBB-Caucasian*; 406,968 subjects), and the second subset removed all 3rd degree or closer relatives from the previous (*UKBB-Unrelated*; 342,796 subjects).

### 2 Aligning Rare Diseases and Their Symptoms to Structured Medical Data

In order to analyze rare disease phenotypes at the population scale, we needed to automatically identify the presence/absence of their associated symptoms within individual medical records. Based on previous work [13, 14, 15, 16], we integrated multiple biomedical ontologies/terminologies (see below for details) to map diseases and their symptoms to structured medical data (i.e. diagnostic billing codes). The details concerning this integration process are provided below.

#### 2.1 Selection of Rare Diseases for Cryptic Phenotype Analysis

We selected the set of rare diseases used for latent phenotype analysis using a combination of ontology integration and manual curation. First, we used the Human Disease Ontology[7] [17] to obtain mappings between the Online Mendelian Inheritance in Man (OMIM) database [18] and the ICD terminologies. Building on previous work [19], we then manually curated the OMIM-to-ICD10-CM alignments, selecting (and grouping if necessary) ICD10-CM codes that reliably mapped to a single or relatively homogenous set of OMIM diseases. By integrating this manually curated set with the diseases annotated by the Human Phenotype

[6]See `https://biobank.ctsu.ox.ac.uk/crystal/label.cgi?id=2002` for details regarding diagnostic information available within the UKBB

[7]`http://www.obofoundry.org/ontology/doid.html`—

Ontology[8] (HPO), we generated a raw list of 166 rare disease phenotypes aligned to both the ICD10-CM and HPO terminologies.

From this set of 166 rare diseases, we further filtered them according to:

1. Their prevalence within the UCSF-CDW
2. Their number of annotated HPO terms

More specifically, we included all diseases for downstream analyses that had a population prevalence of at least $5 \times 10^{-5}$. In addition, we limited our analyses to those diseases with at least 5 annotated HPO terms, as our model for latent phenotype inference showed instability for diseases annotated with less HPO terms (see below). The final set of included diseases (along with modes of inheritance, prevalence and assigned ICD10/ICD10-CM codes) is provided as `SupplementalDataFile_1.txt`.

#### 2.2 Alignment of the Human Phenotype Ontology to the ICD10-CM Terminology

We aligned the Human Phenotype Ontology[9] (HPO) [20] to the ICD10-CM terminology [7] in an automated fashion using multiple sources of data. First, we downloaded the HPO-to-ICD10 and HPO-to-ICD9 mappings (obtained through lexical matching using the LOOM tool [21]) from National Center for Biomedical Ontology's BioPortal[10] [22]. We added to these strict lexical matches by integrating the mappings obtained from two other ontologies: the UMLS Metathesaurus[11] [23] and the SNOMED-CT-to-ICD10 mappings obtained through the US Edition of SNOMED[12] [24]. To further expand the final set of HPO-to-ICD10-CM matches, we also downloaded an ICD9-to-ICD10 map from the National Bureau for Economics Research[13], enabling us to generate HPO-to-ICD10-CM mappings transitively through the ICD9 encoding. Finally, similar to [25], we used a partial logical expansion strategy to increase the coverage of our HPO-to-ICD10-CM mappings. More specifically, for cases in which an HPO term had no ICD10 annotations, we aligned it with the ICD10 annotations of its parent as long as the following criteria were met:

1. A child term has only a single parent term
2. The parent term only has a single child term

Overall, the integration of these different datasets plus post-processing enabled the successful alignment of 3,166 HPO terms (total number of terms in the HPO version downloaded on 8/8/2019: 14,707) and yielded 4,922 HPO-to-ICD10 alignments. We further the filtered this list of aligned HPO terms by:

1. Removing terms that were aligned to the ICD10-CM code for a rare disease of interest
2. Removing terms aligned to a disallowed ICD-10-CM code
3. Collapsing two or more HPO terms that aligned to the same ICD-10-CM code (or a strict subset codes)

This reduced the number of unique HPO terms (or combinations of terms) to 1,674. The final HPO-to-ICD10-CM alignments used in this study are provided in `SupplementalDataFile_2.txt`.

We used the phenotype annotations available through the HPO website[14] to assign symptoms (in the form of HPO terms) to the rare diseases of interest (see Supplemental Figure 1 to see a global overview of the symptom-to-disease mappings). Although we did not manually review the assignments, we did evaluate their performance on a simple rare disease diagnosis prediction task. More specifically, ElasticNet classifiers (implemented in sklearn [26]) for rare disease diagnoses were constructed from our training datasets using

[8] `https://hpo.jax.org/app/download/ontology`
[9] `https://hpo.jax.org/app/download/ontology`
[10] `https://bioportal.bioontology.org/`
[11] `https://www.nlm.nih.gov/research/umls/knowledge_sources/metathesaurus/index.html`
[12] File: `tls_Icd10cmHumanReadableMap_US1000124_20190301.tsv`; `https://www.nlm.nih.gov/healthit/snomedct/us_edition.html`
[13] `https://www.nber.org/data/icd9-icd-10-cm-and-pcs-crosswalk-general-equivalence-mapping.html`
[14] https://hpo.jax.org/app/download/annotation

three sets of features: annotated HPO terms, all HPO terms, and the full ICD10 codebook. The classifiers built using the annotated HPO terms performed substantially better than random (assessed using the average precision score [27]) for nearly all of the included disorders (Supplemental Figure 2A). Moreover, although the classifiers constructed using the full ICD10/HPO codebooks performed systematically better on average (Supplemental Figure 2B), the models constructed using the annotated HPO terms were statistically indistinguishable (based on overlapping 95% confidence intervals) from those constructed using the complete ICD10 codebook (which includes >10,000 features) for 39 of 50 disorders (78%, see Supplemental Table 1).

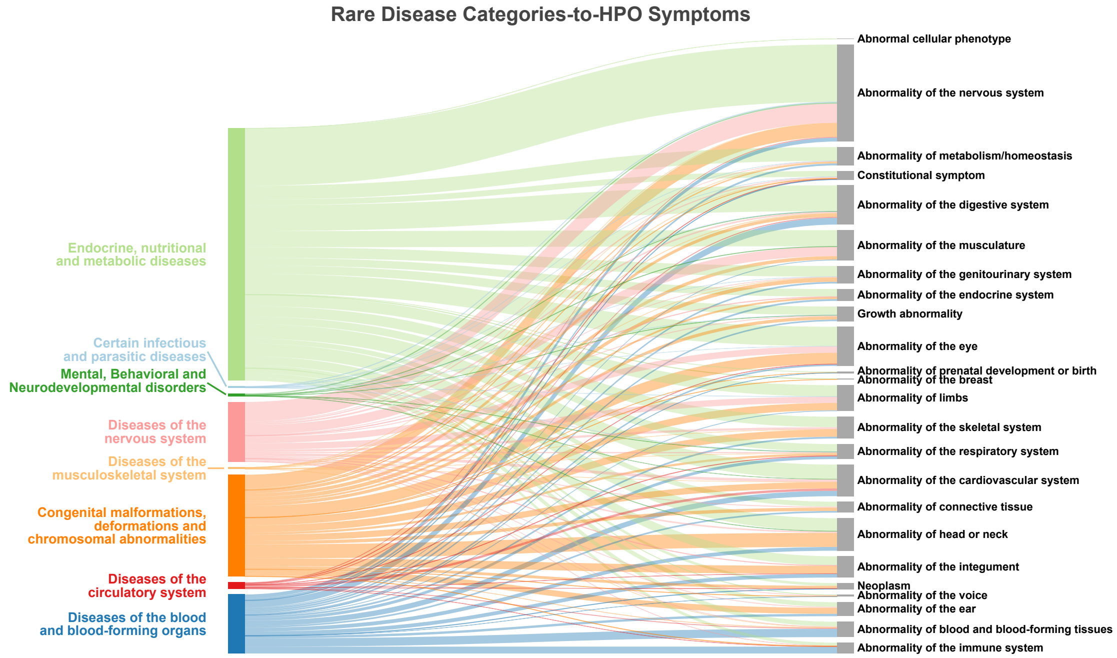


Supplemental Figure 1: A Sankey diagram depicting the alignments between 166 rare, Mendelian diseases (left side) and their HPO-term encoded symptoms (right side). On the left, the Mendelian diseases were grouped according to their parent ICD10-CM chapters such that each colored bar indicates a distinct grouping of disorders, and the height of the bar indicates the total number of diseases in each group. Similarly, on the right side, the HPO symptoms were grouped according to their most general ancestor in the ontology. Annotations between the groups of diseases and symptoms are represented as curved lines of various thickness, with the width of each line proportional to the total number of annotations linking the two groups.

### 3 Cryptic Phenotype Analysis

Our approach to identifying modifiers of Mendelian disease severity relies on the detection of a quantitative but latent (or cryptic) spectrum of variation that drives the observed symptoms of the disorder (i.e. its morbidity or expressivity). However, there are no guarantees that such a spectrum exists. Therefore, we utilized a two-stage process to identify Mendelian diseases that may benefit from this spectrum-based approach. First, we performed statistical inference to estimate the latent phenotypes underlying a set of observed symptoms (see Supplemental Figure 5, left side) in two distinct datasets (UCSF and UKBB). Then, we performed extensive evaluations of the inferred phenotypes to ensure that they: 1) captured the severity of the Mendelian disease of interest, and 2) were consistent across datasets. Cryptic Phenotype Analysis (CPA) refers to the complete two-stage process.

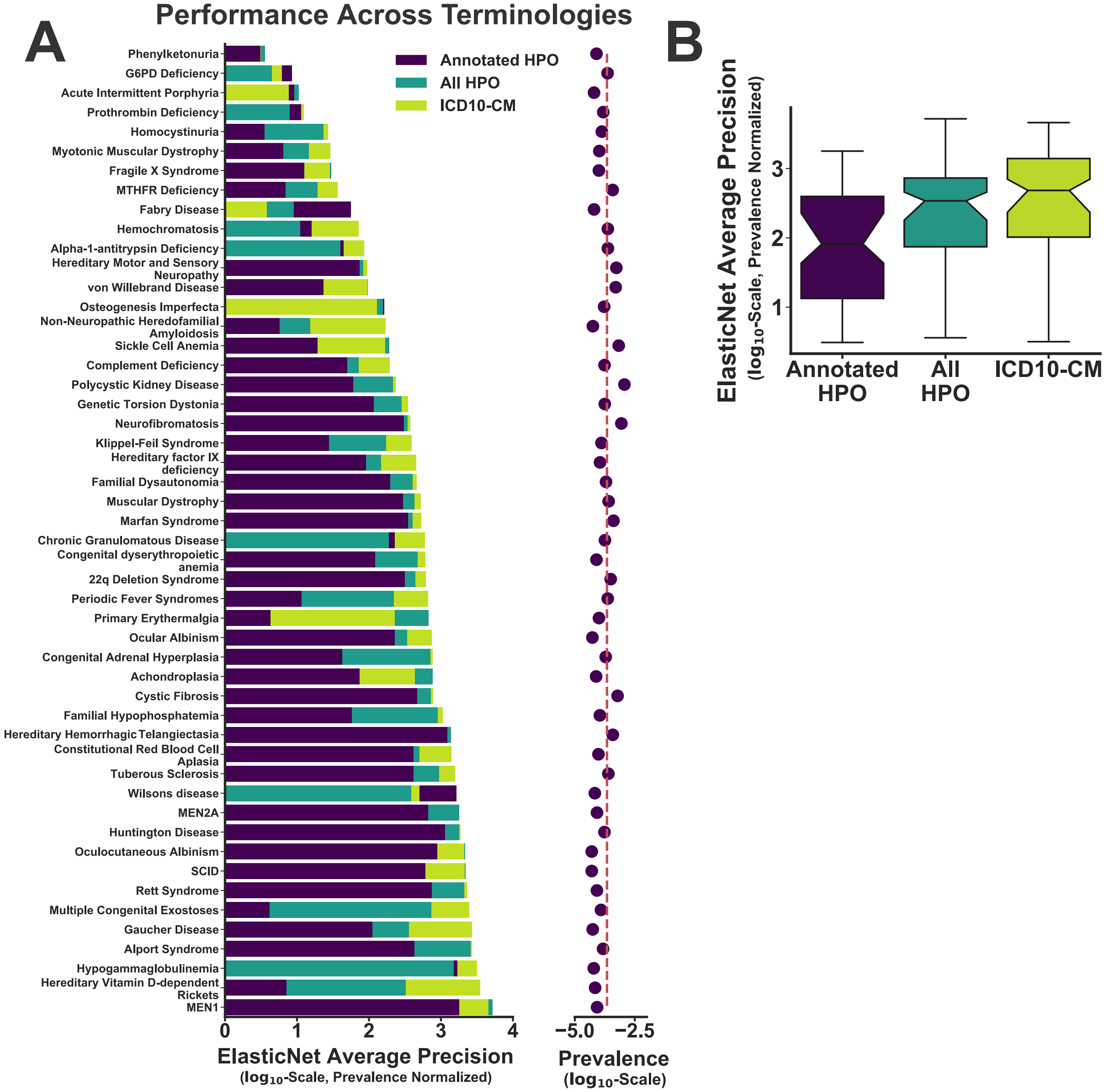


Supplemental Figure 2: Classifiers to predict rare disease diagnoses were constructed from three different terminologies: annotated HPO terms, all HPO terms, and all ICD10-CM codes. A: The average precision score (evaluated in the testing subset) for the three different classifiers is compared across all 50 rare diseases included in the study. The average precision score is normalized by the prevalence of each diagnosis in the dataset, which approximates the average precision score of a random classifier [28]. For reference, the prevalence of each diagnosis is plotted to the right (red line denotes the mean prevalence across all diseases). All classifiers were constructed using the `LogisticRegression` function implemented in sklearn[26], invoking the `elasticnet` penalty and the following hyper-parameter settings: `max_iter=500`, `tol=0.001`, `solver='saga'`, `C=1.0`, and `l1_ratio=0.5`. B: Boxplots comparing the average precision scores across the 3 different terminologies for the 50 diseases included in this study.

Supplemental Table 1: Disease diagnosis classifier performance: annotated HPO vs. ICD10-CM.

| Name | Annotated Avg. Precision (95% CI) | ICD10 Avg. Precision (95% CI) |
|---|---|---|
| Achondroplasia | 0.0057 (0.00041,0.046) | 0.034 (0.0028,0.14) |
| Fabry Disease | 0.0035 (8.9e-05,0.033) | 0.00024 (9.6e-05,0.00089) |
| Gaucher Disease | 0.0063 (0.00071,0.029) | 0.15 (0.0047,0.35) |
| Chronic Granulomatous Disease | 0.041 (0.0077,0.11) | 0.11 (0.04,0.21) |
| Complement Deficiency | 0.0086 (0.0029,0.029) | 0.034 (0.0038,0.096) |
| Hypogammaglobulinemia | 0.1 (0.0055,0.27) | 0.2 (0.045,0.4) |
| Cystic Fibrosis | 0.28 (0.21,0.36) | 0.47 (0.4,0.55) |
| Non-Neuropathic Heredofamilial Amyloidosis | 0.00032 (0.00013,0.0008) | 0.0096 (0.0012,0.046) |
| Homocystinuria | 0.00047 (0.00028,0.00094) | 0.0036 (0.00088,0.019) |
| MTHFR Deficiency | 0.0026 (0.0018,0.0051) | 0.014 (0.003,0.038) |
| Acute Intermittent Porphyria | 0.00058 (0.00013,0.0025) | 0.00048 (0.00013,0.0023) |
| Wilsons disease | 0.11 (0.0046,0.26) | 0.034 (0.007,0.13) |
| Hemochromatosis | 0.0038 (0.0016,0.012) | 0.017 (0.0086,0.038) |
| Hereditary Vitamin D-dependent Rickets | 0.0005 (0.00011,0.0034) | 0.24 (0.072,0.43) |
| Familial Hypophosphatemia | 0.0064 (0.00086,0.038) | 0.12 (0.035,0.27) |
| Alpha-1-antitrypsin Deficiency | 0.011 (0.0044,0.029) | 0.021 (0.008,0.053) |
| Constitutional Red Blood Cell Aplasia | 0.04 (0.015,0.095) | 0.13 (0.044,0.31) |
| Phenylketonuria | 0.00025 (0.00014,0.00044) | 0.00025 (0.00015,0.00053) |
| Ocular Albinism | 0.012 (0.0019,0.052) | 0.04 (0.0021,0.19) |
| Oculocutaneous Albinism | 0.045 (0.0055,0.16) | 0.11 (0.0099,0.28) |
| Genetic Torsion Dystonia | 0.021 (0.005,0.075) | 0.061 (0.024,0.15) |
| Primary Erythermalgia | 0.00043 (0.00026,0.00078) | 0.023 (0.0051,0.084) |
| Fragile X Syndrome | 0.0013 (0.00029,0.0048) | 0.0029 (0.00039,0.022) |
| G6PD Deficiency | 0.002 (0.00074,0.011) | 0.0014 (0.00065,0.0034) |
| Hereditary factor IX deficiency | 0.01 (0.0008,0.064) | 0.051 (0.013,0.15) |
| von Willebrand Disease | 0.012 (0.0054,0.03) | 0.048 (0.024,0.088) |
| Prothrombin Deficiency | 0.0017 (0.0003,0.012) | 0.0019 (0.00084,0.0043) |
| Hereditary Hemorrhagic Telangiectasia | 0.47 (0.37,0.56) | 0.53 (0.44,0.62) |
| Muscular Dystrophy | 0.077 (0.032,0.15) | 0.14 (0.061,0.23) |
| Myotonic Muscular Dystrophy | 0.00067 (0.00016,0.0041) | 0.003 (0.00023,0.023) |
| Hereditary Motor and Sensory Neuropathy | 0.04 (0.026,0.065) | 0.051 (0.031,0.089) |
| Huntington Disease | 0.2 (0.087,0.33) | 0.32 (0.19,0.46) |
| Congenital Adrenal Hyperplasia | 0.0082 (0.0023,0.021) | 0.15 (0.067,0.25) |
| Familial Dysautonomia | 0.039 (0.0083,0.097) | 0.092 (0.036,0.19) |
| Marfan Syndrome | 0.14 (0.08,0.22) | 0.22 (0.14,0.3) |
| Alport Syndrome | 0.064 (0.028,0.14) | 0.41 (0.26,0.55) |
| MEN1 | 0.15 (0.049,0.31) | 0.39 (0.19,0.61) |
| MEN2A | 0.056 (0.01,0.18) | 0.15 (0.033,0.32) |
| Neurofibromatosis | 0.27 (0.21,0.33) | 0.33 (0.27,0.39) |
| Tuberous Sclerosis | 0.1 (0.042,0.18) | 0.39 (0.28,0.5) |
| Multiple Congenital Exostoses | 0.00051 (0.00021,0.0024) | 0.31 (0.16,0.47) |
| Rett Syndrome | 0.062 (0.01,0.17) | 0.19 (0.042,0.36) |
| 22q Deletion Syndrome | 0.098 (0.058,0.18) | 0.19 (0.12,0.28) |
| Polycystic Kidney Disease | 0.07 (0.051,0.099) | 0.27 (0.23,0.32) |
| Osteogenesis Imperfecta | 0.027 (0.00097,0.081) | 0.022 (0.0035,0.084) |
| SCID | 0.031 (0.0044,0.14) | 0.11 (0.01,0.3) |
| Sickle Cell Anemia | 0.013 (0.0061,0.031) | 0.11 (0.071,0.16) |
| Klippel-Feil Syndrome | 0.0036 (0.0006,0.02) | 0.051 (0.0081,0.13) |
| Periodic Fever Syndromes | 0.0027 (0.0014,0.0061) | 0.16 (0.087,0.27) |
| Congenital dyserythropoietic anemia | 0.0097 (0.0028,0.034) | 0.048 (0.01,0.15) |

#### 3.1 A Generative Model for Disease Symptoms

Consider the set of $K$ symptoms that are associated with some Mendelian disease of interest. We assume that all of symptoms are binary (present/absent) and permanent (i.e. once diagnosed, they do not resolve). Let $S_{i,j}$ denote the status of the $j$th symptom in the $i$th subject such that $S_{i,j} = 1$ indicates that the patient has received this diagnosis. Furthermore, let $\mathbf{S}$ denote an $N \times K$-dimensional matrix of symptom diagnoses such that the $i$th row of the matrix (denoted $\mathbf{S}_i$) contains the diagnoses for subject $i$, and let $\mathbf{Z}$ denote an $N \times L$ matrix of latent phenotype values, where each column of the matrix represents the magnitude (i.e. severity) of a distinct latent phenotype. We assume that the binary symptom matrix is generated stochastically from the latent phenotypes according to:

$$P(\mathbf{S}|\mathbf{Z}, \theta) = f(\mathbf{Z}; \theta),$$

where $f(\mathbf{Z}; \theta)$ is a function (defined by the parameters $\theta$) that maps the latent phenotypes onto a matrix of symptom probabilities (i.e. $f(\mathbf{Z}; \theta) \in [0, 1]^{N \times K}$). We construct a joint model for $\mathbf{S}$ and $\mathbf{Z}$ by specifying a generative probability distribution for the latent phenotypes themselves:

$$P(\mathbf{S}, \mathbf{Z}|\theta) = P(\mathbf{S}|\mathbf{Z}, \theta) \times P(\mathbf{Z}|\phi), \quad (1)$$

where $\phi$ is the set of parameters defining the prior distribution for the latent phenotypes.

To fully define the joint model, we must precisely specify the symptom risk function ($f(\mathbf{Z}; \theta)$) and the prior distribution for $\mathbf{Z}$ ($P(\mathbf{Z}|\phi)$). Starting with the latter, we desire a phenotype model that produces continuous random variables, is computationally tractable, and assumes that the $L$ distinct latent phenotypes are independent of one another. This last assumption is important because it aids in identifiability. Our goal is to isolate latent phenotypes that best explain the symptom variability observed within each Mendelian disease. If our model happens to decompose a set of symptoms into multiple phenotypes, then we want them to be independent of one another in order maximize our chances of isolating a single latent component capable of explaining this variability. Therefore, in the current study, we used the isotropic standard gaussian as the prior distribution for the latent phenotypes:

$$\mathbf{Z} \sim \mathcal{N}(\mathbf{0}, \mathbf{I}) \quad (2)$$

where $\mathbf{I}$ is the $L \times L$-dimensional identity matrix. Note, there is no advantage to relaxing the constant variance assumption implicit in this construction; the symptom risk function can always systematically re-scale the latent phenotype values to allow for non-constant variance across the different dimensions.

With respect to the function $f(\mathbf{Z}; \theta)$, we modeled symptom risks using the following generalized linear model:

$$f(\mathbf{Z}; \theta) \propto \exp\left[\mathbf{B}^T + \mathbf{Z}\mathbf{W}\right] \quad (3)$$

where $\mathbf{B}$ denotes a $K$-dimensional vector of symptom-specific offset terms and $\mathbf{W}$ represents an $L \times K$-dimensional matrix of latent phenotype component weights (i.e. $\theta = \{\mathbf{B}, \mathbf{W}\}$). To simplify the interpretation of the inferred latent phenotypes, we restricted the values of $\mathbf{W}$ to be non-negative such that $w_{l,k} \geq 0, \forall l, k$. This restriction was in invoked for two reasons. First, it ensured that all of the symptoms annotated to a Mendelian disease were positively correlated, which should be generally be true by definition. Second, it aided in the interpretation of the inferred latent phenotypes, as it ensured that individuals with large, positive latent phenotype values had the highest risk of being diagnosed with multiple, rare disease symptoms (i.e. the greatest morbidity). Note, more complicated symptom risk functions are possible (ex: multi-layered non-linear neural network), but in our preliminary analyses, such functions were associated with substantially increased complexity while adding little in terms of expressivity. Further research is required to determine if/when more complicated risk functions are necessary to capture the correlation structure among disease symptoms.

#### 3.2 A Variational Approach to Model Inference

Ultimately, we were interested in producing estimates for the latent phenotype matrix (denoted $\hat{\mathbf{Z}}$) given some observed symptom matrix (denoted $\mathbf{S} = \mathbf{s}$). This idea of summarizing an observed dataset with a lower-dimensional matrix of latent variables is also known as matrix factorization (or factor analysis), and there are multiple examples of its successful application to a diverse set of problems in computation biology and genetics [29, 30, 31, 32]. Although many general-purpose matrix factorization algorithms have been developed, most were designed for continuously-distributed observed data and/or do not impose independence assumptions among the latent variables. Therefore, we developed an approximate, variational Bayesian inference algorithm that is specific to the model outlined in Equation 1 using an approach that has become popular within the machine learning community due to its scalability and overall performance [33, 34].

More specifically, our goal was to infer a posterior distribution over $\mathbf{Z}$ given the observed symptom matrix $\mathbf{S} = \mathbf{s}$ and the parameters $\theta$ (i.e. $P(\mathbf{Z}|\mathbf{s}, \theta)$). To do so, we first defined the likelihood of the observed data for our model:

$$P(\mathbf{s}|\mathbf{Z}, \theta) = \prod_{i=1}^{N} \prod_{j=1}^{K} \left[f(\mathbf{Z}; \theta)_{i,j}\right]^{s_{i,j}} \times \left[1 - f(\mathbf{Z}; \theta)_{i,j}\right]^{1-s_{i,j}},$$

where $f(\mathbf{Z}; \theta)_{i,j}$ denotes element-$(i, j)$ of the matrix produced by the symptom risk function. The joint likelihood for the data and the latent phenotypes is:

$$P(\mathbf{s}, \mathbf{Z}|\theta) = P(\mathbf{s}|\mathbf{Z}, \theta) \times P(\mathbf{Z}),$$

and to specify the desired posterior distribution, we need to compute the following marginal likelihood (also called the model evidence):

$$P(\mathbf{s}|\theta) = \int P(\mathbf{s}, \mathbf{Z}|\theta) d\mathbf{Z}.$$

Because this integral is analytically intractable, we produced a lower bound approximation to the model evidence (denoted ELBO) by invoking Jensen's Inequality:

$$\begin{aligned} \log P(\mathbf{s}|\theta) &\geq \int q(\mathbf{Z}|\mathbf{s}, \psi) \log \left[\frac{P(\mathbf{s}, \mathbf{Z}|\theta)}{q(\mathbf{Z}|\mathbf{s}, \psi)}\right] \\ &= \mathbb{E}_{q(\mathbf{Z}|\mathbf{s}, \psi)}\left[P(\mathbf{s}|\mathbf{Z}, \theta)\right] + KL\left[q(\mathbf{Z}|\mathbf{s}, \psi)||p(\mathbf{Z})\right] \\ &\equiv \mathcal{L}_{\theta, \psi} \end{aligned} \tag{4}$$

where $q(\mathbf{Z}|\mathbf{s}, \psi)$ denotes an approximate posterior distribution over $\mathbf{Z}$ defined by the parameter set $\psi$, $\mathbb{E}_G\left[F\right]$ denotes the expectation of $F$ with respect to $G$, and $KL\left[F||G\right]$ denotes the Kullback-Leibler (KL)-divergence from $G$ to $F$. By optimizing this lower bound with respect $\theta$ and $\psi$, we obtain an estimate of the model marginal likelihood (i.e. the evidence) while simultaneously learning an approximate posterior distribution over the latent phenotypes. Further details regarding the general variational approximation and its application to statistical models can be found in [35].

In order for the lower bound defined in Equation 4 to be practically useful, we need specify the approximate posterior distribution $q(\mathbf{Z}|\mathbf{s}, \psi)$. One approach to defining this function is to invoke the mean field approximation, which restricts the joint posterior distribution over the latent variables to be conditionally independent given a set of variational parameters (i.e. $q(\mathbf{Z}|\mathbf{s}, \psi) = \prod_i q(\mathbf{Z}_i|\mathbf{s}_i, \psi_i)$). Although the optimal solution for $q(\mathbf{Z}|\mathbf{s}, \psi)$ under the mean field approximation can sometimes be derived analytically [35], this is not true for the model defined in Equation 1. Therefore, we used the approach outlined in [33, 34] and directly set $q(\mathbf{Z}_i|\mathbf{s}, \psi)$ to have a specific functional form: a multi-layered non-linear neural network[15].

There are several advantages to this approach. First, instead of optimizing a unique set of variational parameters for each observed datapoint (as is the case for the mean field approximation), the posterior distribution for every datapoint arises from a deterministic function that relies on the same set of parameters. This is known as amortized inference [34], and it enables variational inference algorithms to scale to very large datasets. Second, optimization problems involving non-linear neural networks have become common in the machine learning community [36], and multiple, highly-flexible software libraries now exist that enable the rapid development of scalable optimization algorithms for non-linear neural networks [37] and complex probability models [38].

[15]Specifically, we utilized a neural network with 2 hidden layers each containing 64 Rectified Linear Units (ReLU) to allow for non-linearities. Details concerning the implementation can be found in the source code: `https://github.com/daverblair/vlpi`.

##### 3.3 Model Inference in Practice

As discussed above, model inference using the variational approach proceeds by finding the parameter values (denoted $\hat{\theta}$ and $\hat{\psi}$) that maximize the lower bound specified in Equation 4. When a neural network is chosen as the approximate posterior distribution, this objective function is generally maximized using some version of stochastic gradient descent, an iterative algorithm that scales well to large datasets and complex models [36]. Although theoretically guaranteed to converge to a local optimum under certain conditions, the performance of stochastic gradient descent can vary widely across datasets, models, and implementations [36].

To mitigate some of this variability, we aimed to design an inference algorithm that was flexible and enabled excellent *any-time* performance, as we wanted to rapidly deploy our statistical model to many different sets of disease symptoms without having to individually tune the algorithm's hyper-parameters. In our implementation, we utilized an adaptive gradient descent method that includes a penalty term to avoid overfitting[16] (AdamW, see [39]). With respect to the learning rate of the algorithm, we initially started with a very small value, which was increased towards a large, maximum learning rate prior to decreasing the rate down to a small value once again (one-cycle learning rate scheduler). This particular approach has been shown to enable fast and accurate training of complex neural networks [40], and we found this to be true for our model as well.

In some cases, however, we found that different runs of the optimization algorithm could yield substantially different results, particularly with respect to the symptom risk function parameters ($\theta$). This observation appeared to be driven by convergence to local modes in the objective function, likely exacerbated by the stochastic nature of the inference algorithm. To avoid local modes that tended to result in simpler but sub-optimal models, we utilized the annealing approach described in [41] to slowly incorporate the KL-divergence term into the lower bound specified in Equation 4. We found that this annealing approach substantially improved the consistency of the inferred models across different initializations (see below). Finally, by default, we monitored for algorithm convergence by computing the (untempered) ELBO on a held-out sample of data (termed the validation dataset). Unless otherwise noted, we withheld 20% of the training dataset for validation. Note, this validation dataset is distinct from the *testing dataset*, which was only used in the final stage of our cryptic phenotype analysis (see below). The inference algorithm outlined above was implemented using the Pyro Probabilistic Programming Language [38] and PyTorch [37]. Additional details regarding the algorithm, including default hyper-parameters (which were used unless otherwise noted), can here can be found on Github: `https://github.com/daverblair/vlpi`.

##### 3.4 Assessing Model Convergence

The inference algorithm described above was designed in order maximize inferential consistency without having to tune a unique set of hyper-parameters for each symptom dataset. That said, inconsistent inference results were still possible, especially given the stochastic nature of the algorithm coupled with the multimodality of the objective function. Therefore, we wanted to ensure that the model inference results were consistent and robust across multiple model fitting attempts.

To do so, we fit the latent phenotype model to the symptom sets corresponding to each rare disease using 20 different trials of our stochastic inference algorithm. Each trial used the same set of algorithmic hyper-parameters (maximum learning rate: 0.02, mini-batch size: 5000, max. epochs: 2000, convergence error tol.: $10^{-6}$). To systematically compare models across different inference runs, we developed two statistics to assess their similarity. First, we defined a simple measurement that compares the quality of the model fits within our training dataset. More specifically, we computed the per-datum model perplexity for every subject according to:

$$\begin{aligned} \mathcal{P}(\mathbf{s}_i|\theta^{\mathbf{q}}, \psi^{\mathbf{q}}) &= -\int q(\mathbf{Z}_i|\mathbf{s}_i, \psi^{\mathbf{q}}) \log \left[ \frac{P(\mathbf{s}_i, \mathbf{Z}_i|\theta^{\mathbf{q}})}{q(\mathbf{Z}_i|\mathbf{s}, \psi^{\mathbf{q}})} \right] d\mathbf{Z}_i \\ &\geq -\log P(\mathbf{s}_i|\theta), \end{aligned} \tag{5}$$

where the superscript $q$ is used to indicate that these parameters define the model obtained from the $q$th trial. In practice, the integral in Equation 5 cannot be computed analytically, so we computed $\mathcal{P}(\mathbf{s}_i|\theta, \psi)$

[16]Penalty term set to $10^{-4}$ by default.

using a stochastic (Monte Carlo) approximation:

$$\mathcal{P}(\mathbf{s}_i|\theta,\psi) \approx -\sum_{j=1}^{M} \left[ \log P(\mathbf{s}_i, \mathbf{Z}_i^j|\theta) - \log q(\mathbf{Z}_i^j|\mathbf{s}_i,\psi) \right],$$

where $\mathbf{Z^j}$ denotes the $j$th of $M$ random samples obtained from approximate posterior defined by $q(\mathbf{Z}|\mathbf{s},\psi)$. By default, we set $M = 10$; we experimented with larger values of $M$ but found no obvious improvement in performance. To compare the quality of two models, we simply computed the following paired perplexity statistic:

$$\mathcal{P}(\theta^{\mathbf{q}},\psi^{\mathbf{q}}||\theta^{\mathbf{r}},\psi^{\mathbf{r}}) = \sum_{i=1}^{N} \left[ P(\mathbf{s}_i|\theta^{\mathbf{q}},\psi^{\mathbf{q}}) - P(\mathbf{s}_i|\theta^{\mathbf{r}},\psi^{\mathbf{r}}) \right],$$

where $\mathcal{P}(\theta^{\mathbf{q}},\psi^{\mathbf{q}}||\theta^{\mathbf{r}},\psi^{\mathbf{r}}) > 0$ indicates that $q$th model has systematically higher perplexity (i.e. results in a poorer fit) than the $r$th model. Practically speaking, there was a fair amount noise associated with this statistic, so we used bootstrapped resampling [42] to estimate a 95% confidence interval. The top-performing model was selected as the inference trial that resulted in the lowest perplexity after accounting for sampling error (i.e. the 95% CI excluded 0 after multiple testing correction); ties were broken arbitrarily.

The previously described statistic allowed us to select a top performing model while also determining if multiple models were recovered that were of similar quality. However, it does not directly address the issue of multimodality, as we could theoretically recover models of similar quality but with different parameter estimates. To assess parameter similarity, we took advantage of the fact that our linear symptom risk function is simply a matrix of component weights ($\mathbf{W}$), and therefore, models that converged to similar modes in the objective function should have similar risk functions. To assess risk function similarity, we developed a statistic based on the coefficient of determination ($R^2$). First, we aligned the risk matrices from the $q$th and $r$th model (denoted $\mathbf{W}^q$ and $\mathbf{W}^r$) by solving the orthogonal procrustes problem [43], which is necessary because the models have rotational invariance. Then, we computed the similarity between two symptom risk functions as follows. Let $R^2(\mathbf{X},\mathbf{Y})$ denote the coefficient of determination between the two vectors $\mathbf{X}$ and $\mathbf{Y}$. The similarity between two symptom risk matrices was computed according to:

$$\mathcal{R}(\mathbf{W}^q,\mathbf{W}^r) = \sum_{l=1}^{L} \frac{R^2(\mathbf{W}_l^q,\mathbf{W}_l^r)}{w_l} \tag{6}$$

where the subscript $l$ indexes the latent components and $w_l$ is a weighting parameter defined as:

$$w_l = \frac{||\mathbf{W}_l^q|| \times ||\mathbf{W}_l^r||}{\sum_{l=1}^{L} ||\mathbf{W}_l^q|| \times ||\mathbf{W}_l^r||}.$$

The weighting parameters are necessary in order to decrease the contribution from *shadow components* (latent phenotypes whose component weights in the symptom risk function are essentially 0, see below for details), which can lead to an irrelevant degradation in the statistic.

In practice, after identifying the top performing model based on the per-datum perplexity, we then computed the pairwise $R^2$ statistic across all 20 trials (see Supplemental Figure 3A-3C for examples). Models were deemed to be adequately consistent if there was a cluster of at least 3 trials (including the top-performer) with an average $R^2$ measurement $\geq 0.8$. For some diseases (ex: Alpha-1-antitrypsin Defiency, see Supplemental Figure 3A), the algorithm was incredibly consistent and returned nearly identical models across all of the trials. For other diseases (ex: Hereditary Hemorrhagic Telangiectasia, see Supplemental Figure 3B), there was less overall consistency but many of the inferred models were nearly identical. In a handful of cases, the algorithm clearly detected multiple, disparate modes across trials but still met our consistency criteria (ex: von Willenbrand Disease, Supplemental Figure 3C). Overall, the symptom sets for 42 of the 50 diseases (85%) included in our analyses met the consistency criteria within the UCSF dataset using our baseline algorithmic hyper-parameters

For the remaining diseases, manual review revealed that many of them were aligned to a large number of symptoms that were loosely or even entirely unassociated with the disease based on clinical knowledge.

We hypothesized that the inclusion of large numbers of unrelated symptoms was detrimental to inference, as it forced the model to try to account for an unnecessarily complex correlation structure (see below and Supplemental Figure 4). Therefore, we manually curated the symptoms aligned to the 8 diseases that failed our consistency tests and re-ran our inference algorithms. This resulted in the convergence of an additional 4 diseases. However, the symptom sets for 4 diseases still failed to converge (ex: Marfan Syndrome, see Supplemental Figure 3D and 3E for fitting results using the initial and revised symptom sets, respectively). In these cases, we attempted to improve convergence by increasing the maximum learning rate (0.05) and the maximum number of training epochs (4000). On simulated data, we found that such changes to the hyper-parameters often improved the consistency of the model fits by allowing training to occur over a greater number of epochs. In the case of our actual symptom datasets, these changes enabled 1 of the 4 remaining diseases to meet our convergence criteria within the UCSF dataset (Marfan Syndrome, see Supplemental Figure 3D-F).

Note, this same procedure was used to assess model convergence in the UKBB. However, only 38 of the 50 phenotype models converged in this dataset. The discrepancy in convergence is likely multifactorial, and possible causes include the UKBB's smaller sample size, lower number of available features, and differences in demographics/acquisition.

#### 3.5 Estimating Effective Rank

Similar to algorithmic hyper-parameters, the selection of model hyper-parameters (model parameters not optimized during learning) can also impact inference results. With respect to the model defined in Equation 1, there is actually only a single hyper-parameter [17], which is the number of latent phenotypes included into the model (denoted $L$). A simple approach to selecting the number of latent phenotypes would be to set $L = 1$ and isolate a single latent phenotype for each set of rare disease symptoms. In fact, the Phenotype Risk Score defined in [15] exactly corresponds to a particular solution of the Non-Negative Matrix Factorization problem [45] in which the data matrix (in this case, a log-prevalence-transformed symptom matrix) is decomposed into only a single latent factor[18], which may explain some of its success. However, many sets of symptoms are unlikely to be explained by a single latent phenotype, particularly those that cross multiple organ systems. For example, Hereditary Hemorrhagic Telangiectasia (HHT) is a rare, vascular dysplasia characterized by the development of arteriovenous malformations and telangiectasias throughout the body. Due to a propensity for these malformations to rupture, individuals with the disorder are at increased risk for a variety of related symptoms including epistaxis and GI hemorrhage. However, these same symptoms are observed in patients with, for example, portal hypertension and coagulopathy secondary to cirrhotic liver disease [46]. Therefore, attributing all of the correlation structure among the symptoms of HHT to a single underlying latent phenotype could result in noisy and/or biased inferences.

Somewhat surprisingly, the optimization algorithm outlined above is able to automatically select the appropriate number of latent phenotypes by dropping unnecessary latent components from the symptom risk function (somewhat akin to automatic relevance determination). In other words, when the $l$th latent phenotype does not add useful information to the model, then the elements of the $l$th row of latent phenotype loading matrix ($\mathbf{W}$) converge to 0 . This automated elimination of unnecessary latent phenotypes is likely the product of two features of the model and inference algorithm: the KL-divergence term in the loss function and the penalty parameter in the adaptive gradient estimator [39]. In practice, however, the rows of $\mathbf{W}$ that map to unnecessary latent phenotypes may converge slowly to zero, and it can be challenging to determine whether a latent phenotype is truly unnecessary by simply visually examining $\mathbf{W}$. Therefore, we developed a formal approach for detecting and removing unnecessary latent phenotypes by estimating an *effective rank* for our inferred latent phenotype models.

Briefly, let $\mathbf{S}_{\text{Log-Odds}}$ denote the log-odds matrix for the observed symptom data after producing estimates

[17] Although this is technically true for the model specified in Equation 1, the neural network defining our approximate posterior distribution also has a set of associated hyper-parameters: the number of nodes per hidden layer and the number of layers. In practice, we found that inference results were robust to different choices for these hyper-parameters as long as the neural network was reasonably expressive.

[18] This is only true when using the KL-divergence loss function and the multiplicative update rules. Details regarding the equivalence are not shown here but can be found in [45].

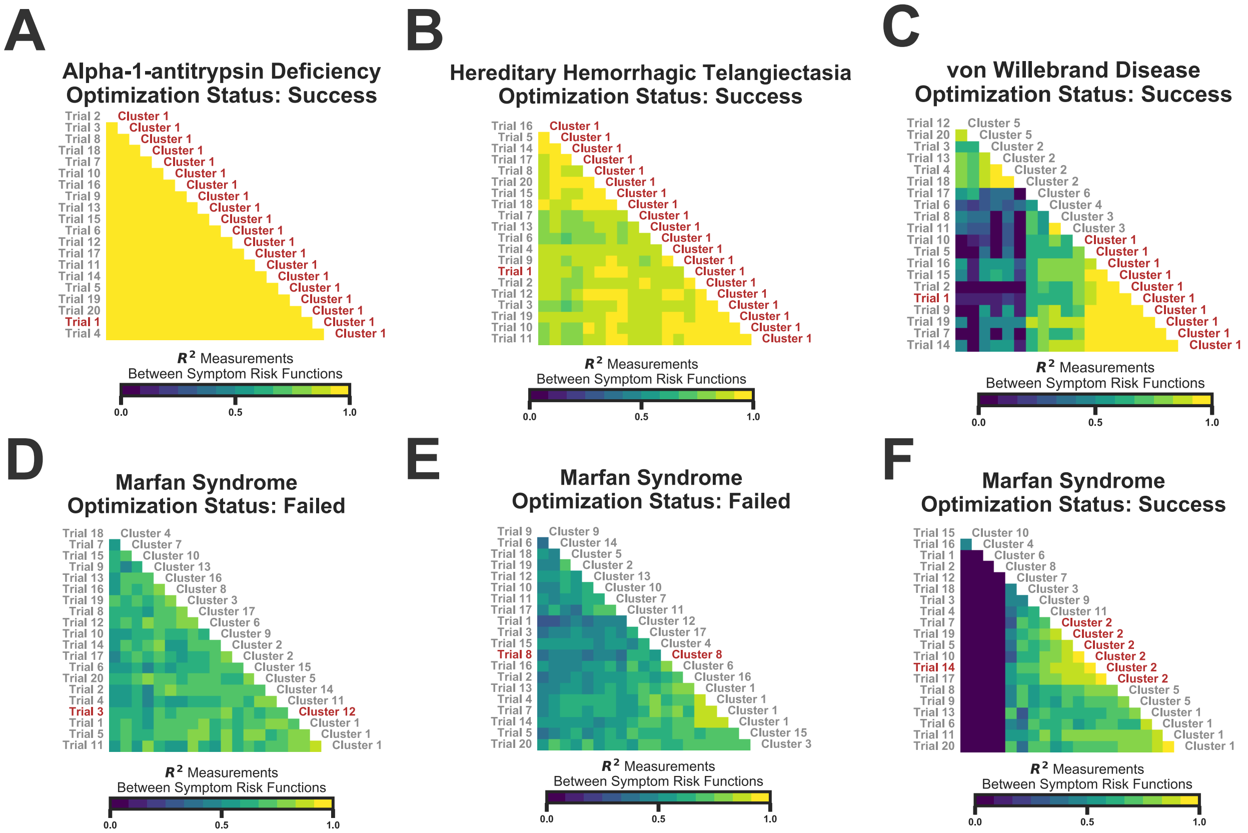


Supplemental Figure 3: Illustrations of our model convergence analysis in the UCSF dataset. In each panel, the heatmap represents the $R^2$ statistic computed across the different model inference trials. The top-performing trial (according to perplexity) is highlighted in red on the left side of each panel, while the cluster containing the top-performing trial is highlighted in red on the right. The pairwise $R^2$ statistics were clustered with the agglomerative method implemented in sklearn [44] using the `average` linkage criterion and a cluster threshold of 0.8. A-C: These panels depict the results for three example diseases that met our convergence criteria using the baseline set of algorithmic hyper-parameters. D-E: These panels depict the convergence results for Marfan Syndrome, which was fit using the baseline set of hyper-parameters (D), the baseline set after pruning unrelated symptoms (E), and our modified set of algorithmic hyper-parameters (F, see text). The models for Marfan Syndrome met our convergence criteria after pruning symptoms and modifying hyper-parameters (F).

of the model parameters (denoted $\hat{\theta}$) and latent phenotypes $\hat{\mathbf{Z}}$:

$$\mathbf{S}_{\text{Log-Odds}} = \hat{\mathbf{Z}}\hat{\mathbf{W}}$$

where $\hat{\mathbf{W}}$ arises directly from the optimization of the lower bound specified in Equation 4 and $\hat{\mathbf{Z}} = \mathbb{E}_{q(\mathbf{Z}|\mathbf{s},\psi)}\big[\mathbf{Z}\big]$. To estimate the effective rank of the model, we first compute the singular value decomposition [47] of the symptom log-odds matrix such that:

$$\mathbf{S}_{\text{Log-Odds}} = \mathbf{U}\mathbf{D}\mathbf{V}^T,$$

where $\mathbf{U}$ and $\mathbf{V}$ denote the left ($N \times K$-dimensional) and right ($K \times K$-dimensional) singular vectors respectively and $\mathbf{D}$ is a $K \times K$-dimensional diagonal matrix of singular values. Importantly, the relative fraction of the variance explained by each of the $K$ singular vectors is given by the following simple equation [47]:

$$\eta_k = \frac{d_k^2}{\sum_{k=1}^{K} d_k^2},$$

where $\mathbf{d}$ is the diagonal of the singular value matrix. We define a model's effect rank (denoted $L_{\mathit{eff}}$) as the number of singular vectors of the symptom log-odds matrix that account for at least $\rho \times 100\%$ of the total variance:

$$L_{\text{eff}} = \sum_{k=1}^{K} \delta(\eta_k \geq \rho),$$

where $\delta(X)$ returns 1 is $X$ is true and 0 otherwise. For our cryptic phenotype models, we set $\rho = 10^{-6}$. On simulated data, we found that $L_{\text{eff}}$ provided an accurate estimate of the number of latent phenotypes underlying a set of observed symptoms.

With respect to the actual diseases, the number of inferred latent phenotypes $L_{\text{eff}}$ was found to be correlated with the number of HPO-annotated symptoms (see Supplemental Figure 4). This isn't entirely surprising, as larger sets of symptoms have a greater chance of capturing multiple pathological processes. This relationship is important to keep in mind when applying latent phenotype models to datasets with large numbers of symptoms, as it can be challenging to consistently infer models with very complex latent phenotypic structures.

#### 3.6 Cryptic Phenotype Identification

Although they generally provide better fits to complex symptom datasets, there are downsides to inferring latent phenotype models with multiple components. With respect to our particular application, we are interested in estimating rare disease morbidity using a single, quantitative latent phenotype inferred from some observed symptom data. When multiple latent phenotypes are inferred, a single latent component must be selected as the top candidate. In some cases, this can be done by simply examining the symptom risk function: the component weights corresponding to the latent phenotype that best captures rare disease morbidity generally closely align with clinical knowledge of symptom frequency. To automate this process, we took advantage of the fact that the rare diseases in our target set (see `SupplementalDataFile_1.text`) were aligned to specific ICD10 codes. These codes, although likely noisy, can serve as proxies for rare disease diagnoses in the population. Therefore, to assign rare diseases to the appropriate cryptic phenotype, we simply identified the latent component in the inferred model that was most predictive of rare disease diagnoses in our training dataset. In other words, the cryptic phenotype was identified as the latent component that generated symptom frequencies (i.e morbidity) that were the most consistent with observed clinical diagnoses.

We produced estimates of the latent phenotypes using the approximate posterior distribution obtained during model inference: $\hat{\mathbf{Z}} = \mathbb{E}_{q(\mathbf{Z}|\mathbf{s},\psi)}\big[\mathbf{Z}\big]$. Following estimation, we sorted the latent phenotypes according to the magnitude of their component symptom weight vectors (i.e. $||\mathbf{W}_l||$ for the $l$th latent phenotype), and we used the top $L_{\text{eff}}$ latent phenotypes as classifiers to predict rare disease diagnoses in our training dataset. The performance of the latent phenotypes at this task was assessed using the average precision score

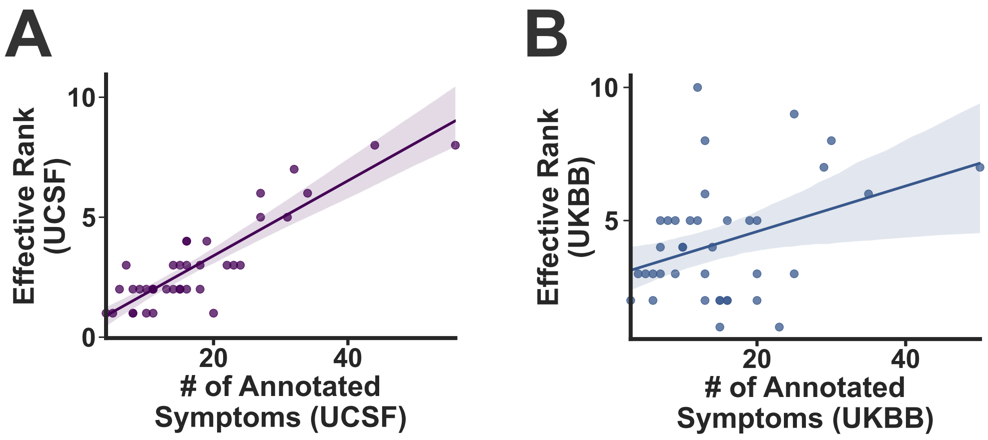


Supplemental Figure 4: For the rare diseases included in this study, the effective rank of the top performing latent phenotype model is plotted against the total number of annotated symptoms. The fraction of variance threshold for determining the effective rank was set to $10^{-6}$. Lines represent the expectation of an ordinary least squares model, and shaded regions indicated the bootstrapped 95% confidence intervals. (A): UCSF Models. (B) UKBB Models.

implemented in sklearn [27]. Importantly, the average precision score is a summary statistic of the precision-recall curve, which generally maintains utility even in the setting of extremely unbalanced datasets (unlike the receiver operating characteristic curve). This approach was used to identify the cryptic phenotypes for actual rare diseases with excellent results (see Main Figures 2 and 3).

##### 3.7 Cryptic Phenotype Evaluation

As discussed in the introduction of this text, not all rare, Mendelian diseases necessarily map to a spectrum of cryptic phenotypic variation, and a variety of artifacts can potentially arise during latent phenotype inference. To mitigate such issues, we took several steps to ensure the cryptic phenotypes inferred using our approach were replicable and consistent. First, cryptic phenotypes were independently inferred in two unique datasets of very different provenance (UCSF and the UKBB). Note, the UKBB is a smaller dataset that is likely depleted for severely ill patients (due to the nature of its recruitment). Therefore, fewer models were successfully inferred in this dataset, and when inference did succeed, the models may be of lower quality. Second, the cryptic phenotypes inferred for each disease were systemically compared across diagnosed cases and controls. If a cryptic phenotype indeed captures Mendelian disease severity, then it should be systematically higher among diagnosed cases. This relationship was assessed by computing bootstrapped estimates of the mean cryptic phenotypes among diagnosed cases and controls [42] with $P$-values computed from the re-sampling statistics. Note, this is only possible for diseases with available ICD10 diagnostic codes. Such information is available for all of the diseases in the UCSF dataset (by design), but some diseases are missing diagnostic codes in the UKBB dataset (see Supplemental Data File 1). When this data is missing, then increased severity among Mendelian disease cases was only assessed in the UCSF dataset. Of the 38 diseases that converged in both datasets, the cryptic phenotypes for 31 were significantly elevated among the diagnosed cases in the UCSF datasets (after correcting for multiple testing).

Now, the above analysis only indicates that independent cryptic phenotypes can be inferred in the UCSF dataset, but it does assess whether the results replicated in the UKBB. To ensure replicability and consistency, we applied the cryptic phenotype models inferred in the UKBB dataset to the UCSF dataset. Note, it is not straightforward to perform the inverse analysis (replicate UCSF models in the UKBB), as the UKBB is missing many of the ICD10-CM codes that are utilized by the UCSF models. After applying the UKBB models to the UCSF data, we assessed replicability and consistency in three ways. First, we ensured that the same latent component was identified as the cryptic phenotype in both datasets (true for 24 out of 31 diseases). Next, we assessed if the cryptic phenotype severity was elevated among Mendelian disease cases using this new model, and we also ensured that this result replicated in the UKBB (when possible, as ICD10 diagnostic codes are not available for all of the disorders in the UKBB). This was true for 18 of the 24 remaining disorders. Finally. we computed the coefficient of determination ($R^2$) between the cryptic phenotypes inferred by the UKBB and UCSF models within the UCSF dataset [19]. There was a fair degree of variability in $R^2$ estimates across diseases. In the end, we selected only those cryptic phenotypes with an $R^2$-value greater than or equal to 0.2 (correlation coefficient=0.4) for downstream analyses (see Supplemental Data File 4). This choice was arbitrary, and we suspect that this low threshold may in part explain our reduced ability to identify common variant modifiers for some of the diseases.

##### 3.8 The Complete Cryptic Phenotype Analysis Pipeline

A summary of our global approach to cryptic phenotype inference is provided in Supplemental Figure 5. Note, much of the complexity associated with this approach stems from the fact that two independent models were inferred in two unique datasets. Ideally, future iterations of this approach would enable joint inference across datasets, which would negate much of this complexity. However, assessing convergence and replicability will always an important part of any model-based approach to phenotype-driven analyses.

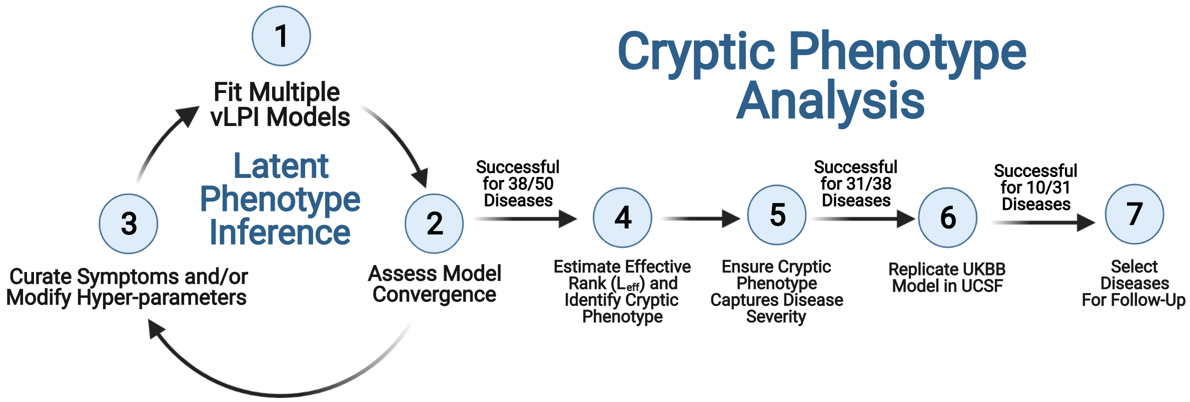


Supplemental Figure 5: Flow diagram for the Cryptic Phenotype Analysis pipeline used in this study. Further details regarding the individual steps can be found in Section 3. This figure was created with BioRender.com.

##### 3.9 Cryptic Phenotype Replication in Arbitrary Datasets

To facilitate the replication of our results in other datasets, we wrote a small software package that imputes the cryptic phenotypes for the five diseases included in our follow up analyses into arbitrary datasets using flat text files containing ICD10-CM/ICD10-UKBB diagnoses. This software package can be found on Github: `https://github.com/daverblair/CrypticPhenoImpute`. This package does not perform any model re-fitting and simply outputs the results of the models that were trained using the UCSF/UKBB datasets. It is of course possible to train new latent phenotype models on existing/independent datasets, but this requires the use of the `vlpi` software package: `https://github.com/daverblair/vlpi`. Additional details concerning the use of both software packages can be found on Github.

[19] It is only possible to perform this analysis in the UCSF dataset due to different encoding used by the UKBB.

Supplemental Table 2: Genome-wide significant common variant loci associated with the cryptic phenotype inferred for A1ATD.

| rsID | Chrom. | Position | Ref. Allele | Min. Allele | $\beta_{SNP}$ (std. err.) | $P$-value |
|---|---|---|---|---|---|---|
| rs1828591 | 4 | 145480780 | A | G | -0.005 (8.21e-04) | 1.51e-09 |
| rs16969968 | 15 | 78882925 | G | A | 0.006 (8.54e-04) | 3.30e-12 |
| rs62206958 | 20 | 62002451 | C | T | 0.009 (1.55e-03) | 2.46e-08 |

Supplemental Table 3: Genome-wide significant common variant loci associated with the cryptic phenotype inferred for AS.

| rsID | Chrom. | Position | Ref. Allele | Min. Allele | $\beta_{SNP}$ (std. err.) | $P$-value |
|---|---|---|---|---|---|---|
| rs7767099 | 6 | 28768698 | C | T | -0.015 (2.06e-03) | 4.31e-13 |
| rs9521732 | 13 | 111033821 | C | A | 0.008 (1.38e-03) | 2.71e-08 |
| rs73045269 | 19 | 41825191 | C | T | -0.010 (1.79e-03) | 8.71e-09 |

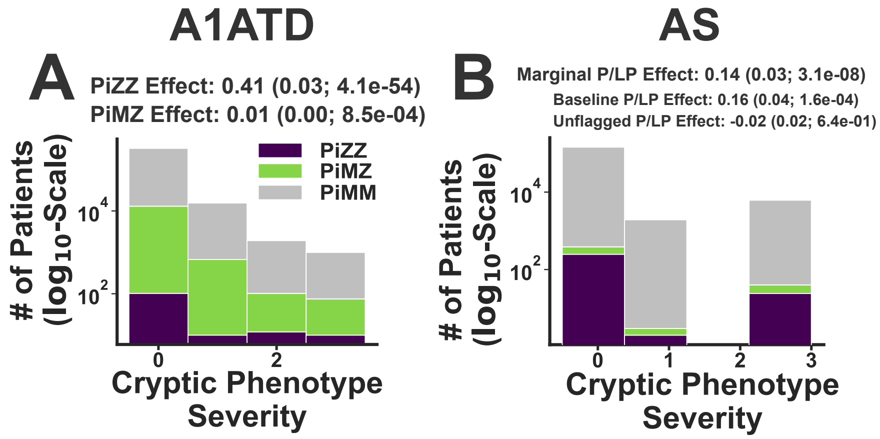


Supplemental Figure 6: Exome sequencing validation of the inferred cryptic phenotypes, continued. (A): The distribution of cryptic phenotype severity is stratified by the three different $\alpha$-1-antitrypsin deficiency (A1ATD) genotypes: Pi*MM (grey), Pi*MZ (green), and Pi*ZZ (purple). Marginal effect sizes for the two pathogenic genotypes are shown at the top. (B): The distribution of cryptic phenotype severity for Alport Syndrome (AS) is stratified by genotype. Reference (grey), P/LP Carrier (purple), and Flagged P/LP Carrier (green). The marginal (flagged and unflagged variants) P/LP cryptic phenotype effect size is shown at the top of the panel. The baseline and unflagged (P/LP x Unflagged) variant effects are displayed below the marginal effect statistics. Note, the parentheses contain the standard errors and $P$-values for the genetic effects, which were estimated using ordinary least squares.

Supplemental Table 4: Genome-wide significant common variant loci associated with the cryptic phenotype inferred for AD-PCKD.

| rsID | Chrom. | Position | Ref. Allele | Min. Allele | $\beta_{SNP}$ (std. err.) | $P$-value |
|---|---|---|---|---|---|---|
| rs12046278 | 1 | 10799577 | T | C | 0.010 (1.22e-03) | 2.64e-15 |
| rs12037987 | 1 | 113042822 | T | C | 0.015 (2.24e-03) | 5.74e-11 |
| rs2586886 | 2 | 26932031 | T | C | 0.010 (1.19e-03) | 4.32e-18 |
| rs1124873 | 3 | 27445430 | G | A | -0.007 (1.20e-03) | 1.35e-09 |
| rs9993149 | 4 | 38404640 | G | T | 0.006 (1.16e-03) | 3.05e-08 |
| rs16998073 | 4 | 81184341 | A | T | 0.013 (1.28e-03) | 4.74e-24 |
| rs17615906 | 5 | 128018413 | T | C | 0.009 (1.56e-03) | 5.44e-09 |
| rs9313772 | 5 | 157804457 | C | T | -0.008 (1.22e-03) | 4.07e-10 |
| rs9375459 | 6 | 127147704 | C | T | 0.009 (1.17e-03) | 2.23e-15 |
| rs117564322 | 7 | 150684021 | G | A | 0.020 (3.30e-03) | 2.24e-09 |
| rs330081 | 8 | 9154065 | A | C | -0.007 (1.17e-03) | 5.07e-09 |
| rs11250099 | 8 | 10818657 | G | A | 0.007 (1.16e-03) | 4.09e-09 |
| rs11014171 | 10 | 18711195 | C | T | -0.007 (1.23e-03) | 1.39e-08 |
| rs12356674 | 10 | 21889083 | G | T | 0.008 (1.29e-03) | 1.86e-09 |
| rs1530440 | 10 | 63524591 | C | T | -0.012 (1.49e-03) | 5.87e-15 |
| rs4980389 | 11 | 1892585 | G | A | 0.008 (1.20e-03) | 3.70e-12 |
| rs35473622 | 11 | 100692566 | A | G | -0.010 (1.34e-03) | 7.57e-13 |
| rs76281384 | 12 | 70421102 | G | C | 0.009 (1.64e-03) | 4.90e-08 |
| rs3184504 | 12 | 111884608 | C | T | 0.007 (1.16e-03) | 3.99e-09 |
| rs2627323 | 15 | 81021383 | C | T | 0.008 (1.17e-03) | 3.16e-12 |
| rs7497304 | 15 | 91429176 | G | T | 0.008 (1.24e-03) | 1.66e-10 |
| rs4293393 | 16 | 20364588 | A | G | -0.011 (1.50e-03) | 1.65e-13 |
| rs34445439 | 17 | 7441756 | C | T | 0.007 (1.27e-03) | 8.20e-09 |
| rs17608766 | 17 | 45013271 | T | C | 0.009 (1.64e-03) | 3.64e-08 |
| rs16948048 | 17 | 47440466 | A | G | 0.007 (1.21e-03) | 1.50e-08 |
| rs9895661 | 17 | 59456589 | T | C | -0.009 (1.55e-03) | 4.09e-08 |
| rs167479 | 19 | 11526765 | G | T | -0.007 (1.16e-03) | 1.38e-10 |
| rs1535067 | 20 | 8607333 | C | T | -0.009 (1.46e-03) | 1.21e-09 |
| rs6078000 | 20 | 10977631 | A | G | 0.008 (1.28e-03) | 8.70e-11 |
| rs16982520 | 20 | 57758720 | A | G | 0.011 (1.78e-03) | 2.91e-09 |

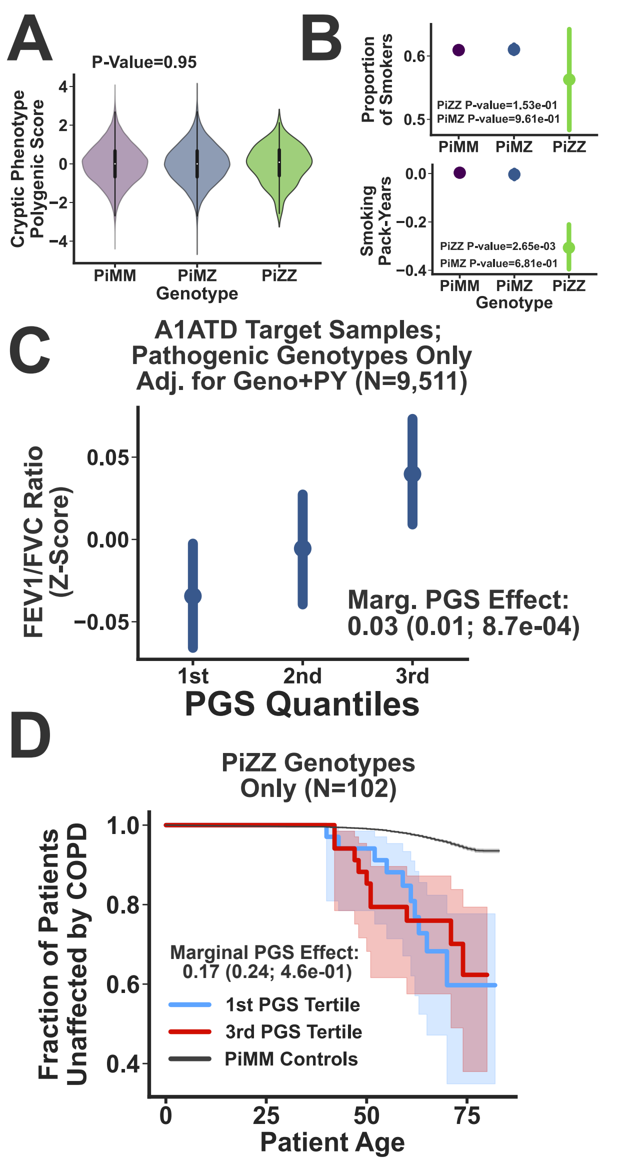


Supplemental Figure 7: Cryptic phenotype-associated genetic variation modifies A1ATD severity, continued. (A): Polygenic load is stratified by A1ATD genotype. Note, there is no association among the genotypes and the PGS scores. (B): Proportion of smokers (top) and reported pack-years (Z-scores, bottom) stratified by genotype. *P*-values summarizing the associations between genotype and smoking history were assessed using logistic (Firth-corrected; LR Test, top) and linear (two-side T-test, bottom) regression. (C): The FEV1/FVC ratio (adjusted for pack-years) is plotted against PGS quantile. This analysis was restricted to pathogenic genotype (Pi*MZ, Pi*ZZ) carriers only. (D): Kaplan-Meier curve for COPD onset, stratified by PGS quantile, is depicted for the Pi*ZZ genotype carriers.

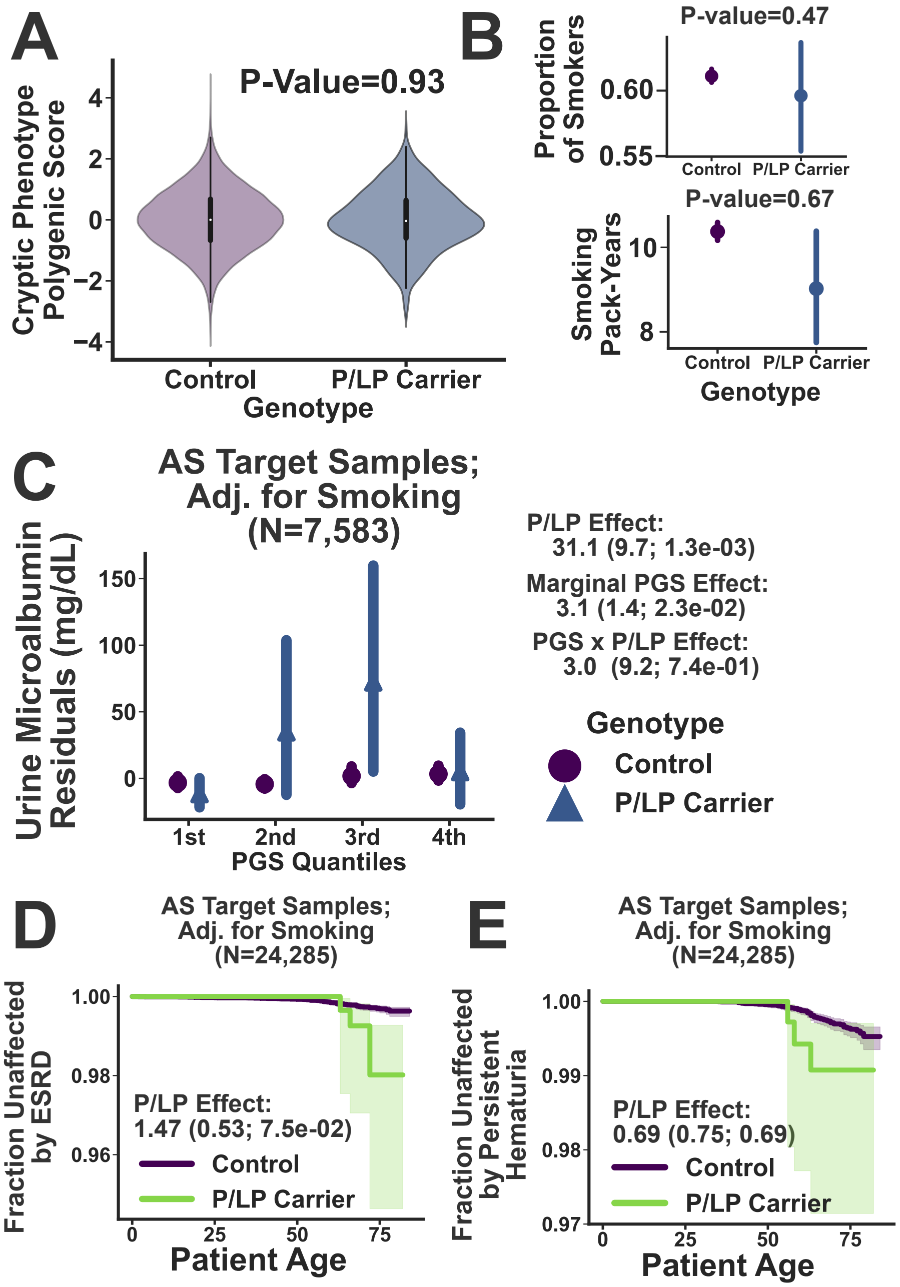


Supplemental Figure 8: Common variation modifies the severity of outcomes associated with Alport Syndrome (AS). (A): Polygenic load is stratified by AS genotype. Note, there is no association among the genotypes and polygenic burden. (B): Proportion of smokers (top) and reported pack-years (bottom) stratified by genotype. *P*-values summarizing the associations between genotype and smoking history were assessed using logistic (Firth-corrected; LR Test) and linear (two-sided T-test) regression. (C): Urine Microalbumin (adjusted for pack-years and baseline covariates) is plotted against PGS quantile and stratified by P/LP genotype. The association statistics for the marginal PGS effect and the genotype-by-PGS interaction effect are depicted to the right. (D, E): Kaplan-Meier curves for ESRD (D) and Persistent Hematuria (E) onset stratified by P/LP carrier status. Association statistics for the P/LP genotype (estimated using Cox-Proportional Hazard regression, see Methods) are displayed within each plot.

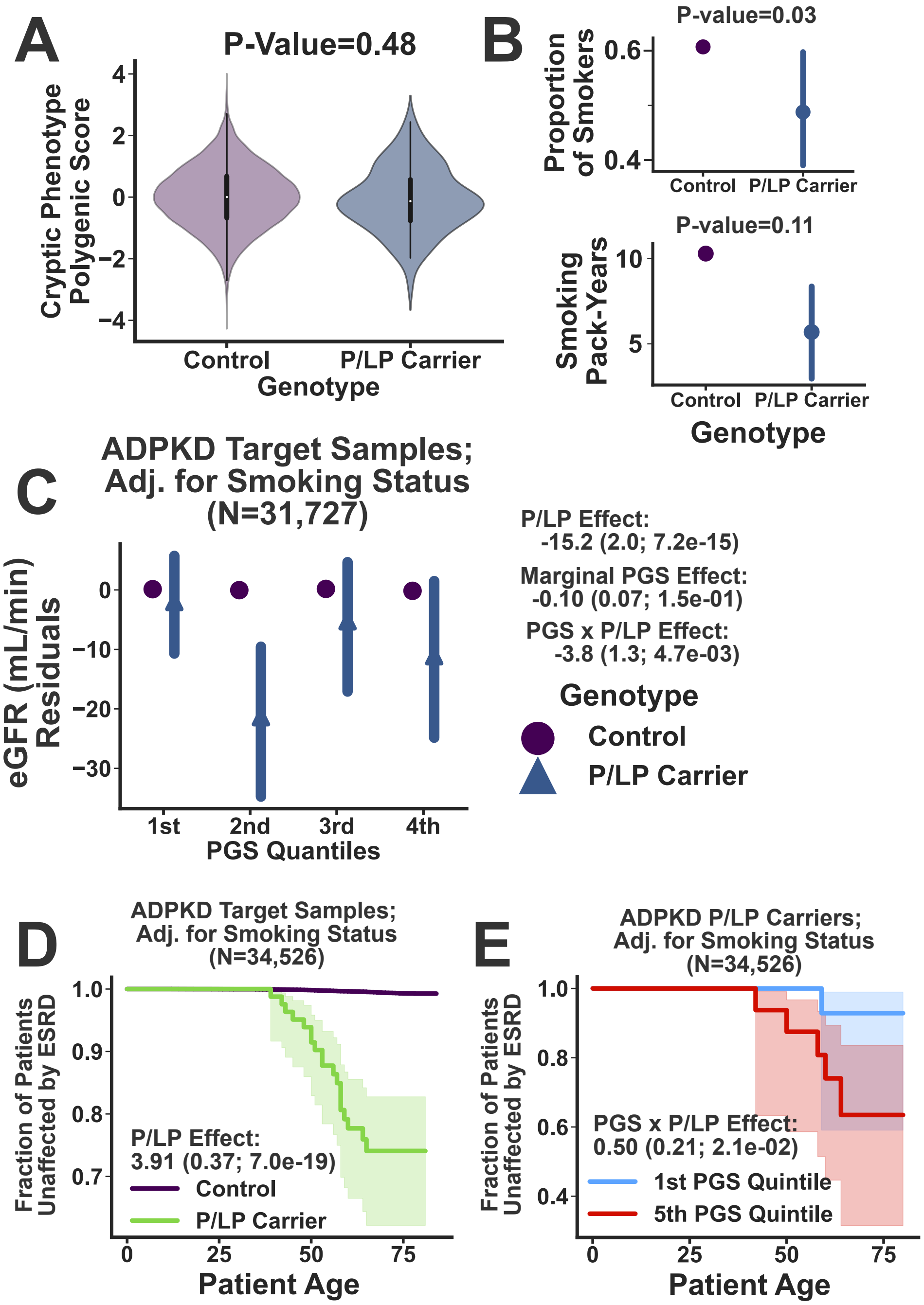


Supplemental Figure 9: Common variation modifies the severity of outcomes associated with Autosomal-Dominant Polycystic Kidney Disease (ADPKD). (A): Polygenic load is stratified by ADPKD genotype. Note, there is no association among the genotypes and polygenic burden. (B): Proportion of smokers (top) and reported pack-years (bottom) stratified by genotype. $P$-values summarizing the associations between genotype and smoking history were assessed using logistic (Firth-corrected; LR Test) and linear (two-side T-test) regression. (C): Estimated Glomerular Filtration Rate (eGFR; adjusted for smoking history and baseline covariates) is plotted against PGS quantile and stratified by P/LP genotype. Association statistics for the marginal PGS effect along with genotype interaction effects are depicted to the right. (D): The Kaplan-Meier curves for ESRD onset is stratified by ADPKD P/LP carrier status. (E) The Kaplan-Meier curve for ESRD onset among P/LP carriers only, stratified by PGS quantile. Association statistics (estimated using Cox-Proportional Hazard regression, see Methods) are displayed within each plot.

[26] Sklearn.linear_model.LogisticRegression — scikit-learn 0.23.1 documentation. https://scikit-learn.org/stable/modules/generated/sklearn.linear_model.LogisticRegression.html.

[27] Sklearn.metrics.average_precision_score — scikit-learn 0.23.1 documentation. https://scikit-learn.org/stable/modules/generated/sklearn.metrics.average_precision_score.html.

[28] Bestgen, Y. (2015). Exact Expected Average Precision of the Random Baseline for System Evaluation. The Prague Bulletin of Mathematical Linguistics *103*, 131–138.

[29] Pritchard, J. K., Stephens, M., and Donnelly, P. (2000). Inference of Population Structure Using Multilocus Genotype Data. Genetics *155*, 945–959.

[30] Brunet, J.-P., Tamayo, P., Golub, T. R., and Mesirov, J. P. (2004). Metagenes and molecular pattern discovery using matrix factorization. Proceedings of the National Academy of Sciences *101*, 4164–4169.

[31] Engelhardt, B. E. and Stephens, M. (2010). Analysis of Population Structure: A Unifying Framework and Novel Methods Based on Sparse Factor Analysis. PLOS Genetics *6*, e1001117.

[32] Lopez, R., Regier, J., Cole, M. B., Jordan, M. I., and Yosef, N. (2018). Deep generative modeling for single-cell transcriptomics. Nature Methods *15*, 1053–1058.

[33] Kingma, D. P. and Welling, M. (2014). Auto-Encoding Variational Bayes. arXiv:1312.6114 [cs, stat].

[34] Kingma, D. P. and Welling, M. (2019). An Introduction to Variational Autoencoders. Foundations and Trends® in Machine Learning *12*, 307–392.

[35] Wainwright, M. J. and Jordan, M. I. (2008). Graphical Models, Exponential Families, and Variational Inference. Foundations and Trends® in Machine Learning *1*, 1–305.

[36] Nielsen, M. A. (2015). Neural Networks and Deep Learning.

[37] Paszke, A., Gross, S., Massa, F., Lerer, A., Bradbury, J., Chanan, G., Killeen, T., Lin, Z., Gimelshein, N., Antiga, L., et al. (2019). PyTorch: An Imperative Style, High-Performance Deep Learning Library. In Wallach, H., Larochelle, H., Beygelzimer, A., d\textquotesingle Alché-Buc, F., Fox, E., and Garnett, R., eds., Advances in Neural Information Processing Systems 32 In Wallach, H., Larochelle, H., Beygelzimer, A., d\textquotesingle Alché-Buc, F., Fox, E., and Garnett, R., eds., Advances in Neural Information Processing Systems 32. (Curran Associates, Inc.).

[38] Bingham, E., Chen, J. P., Jankowiak, M., Obermeyer, F., Pradhan, N., Karaletsos, T., Singh, R., Szerlip, P., Horsfall, P., and Goodman, N. D. (2019). Pyro: Deep Universal Probabilistic Programming. Journal of Machine Learning Research *20*, 1–6.

[39] Loshchilov, I. and Hutter, F. (2019). Decoupled Weight Decay Regularization. arXiv:1711.05101 [cs, math].

[40] Smith, L. N. and Topin, N. (2018). Super-Convergence: Very Fast Training of Neural Networks Using Large Learning Rates. arXiv:1708.07120 [cs, stat].

[41] Sønderby, C. K., Raiko, T., Maaløe, L., Sønderby, S. K., and Winther, O. (2016). How to Train Deep Variational Autoencoders and Probabilistic Ladder Networks. /paper/How-to-Train-Deep-Variational-Autoencoders-and-S%C3%B8nderby-Raiko/2ad303a88ba4fe57a4a3618aa42be90335481acc.

[42] Efron, B. and Tibshirani, R. J. (1993). An Introduction to the Bootstrap. (New York: Chapman and Hall/CRC) 1 edition edition.

[43] Schönemann, P. H. (1966). A generalized solution of the orthogonal procrustes problem. Psychometrika *31*, 1–10.

[44] Sklearn.cluster.AgglomerativeClustering — scikit-learn 0.23.1 documentation. https://scikit-learn.org/stable/modules/generated/sklearn.cluster.AgglomerativeClustering.html.

[45] Lee, D. D. and Seung, H. S. (2001). Algorithms for Non-negative Matrix Factorization. In Leen, T. K., Dietterich, T. G., and Tresp, V., eds., Advances in Neural Information Processing Systems 13 In Leen, T. K., Dietterich, T. G., and Tresp, V., eds., Advances in Neural Information Processing Systems 13. (MIT Press).

[46] McDonald, J. and Pyeritz, R. E. (1993). Hereditary Hemorrhagic Telangiectasia. In Adam, M. P., Ardinger, H. H., Pagon, R. A., Wallace, S. E., Bean, L. J., Stephens, K., and Amemiya, A., eds., GeneReviews®In Adam, M. P., Ardinger, H. H., Pagon, R. A., Wallace, S. E., Bean, L. J., Stephens, K., and Amemiya, A., eds., GeneReviews®. (Seattle (WA): University of Washington, Seattle).

[47] Wall, M. E., Rechtsteiner, A., and Rocha, L. M. (2003). Singular Value Decomposition and Principal Component Analysis. In Berrar, D. P., Dubitzky, W., and Granzow, M., eds., A Practical Approach to Microarray Data Analysis In Berrar, D. P., Dubitzky, W., and Granzow, M., eds., A Practical Approach to Microarray Data Analysis. (Boston, MA: Springer US).
